# Scaling a digital platform for informing and improving child and adolescent mental health service delivery and research: results from the first 10,000 families engaged with myHealthE

**DOI:** 10.64898/2026.01.23.26344624

**Authors:** Alice Wickersham, Craig Colling, Sarjhana Ragunathan Brindha, Jonathan Hind, Jane Anderson, Rosemary Sedgwick, Stephanie J Lewis, Anna Morris, Edmund Sonuga-Barke, Kasia Kostyrka-Allchorne, Claire Ballard, Richard Dobson, Robert Stewart, Matthew Hotopf, Emily Simonoff, Lukasz Zalewski, Jessica Penhallow, Johnny Downs, the CAMHS Digital Lab

**Affiliations:** CAMHS Digital Lab, King’s Maudsley Partnership, Institute of Psychiatry, Psychology & Neuroscience (IOPPN), King’s College London, London, UK; South London and Maudsley NHS Foundation Trust, London, UK; Department of Child and Adolescent Psychiatry, IOPPN, King’s College London, UK; Division of Psychiatry, University College London, UK; Department of Psychology, School of Biological and Behavioural Sciences, Queen Mary University of London, London, UK; Care for Long Term Conditions Division, Florence Nightingale Faculty of Nursing, Midwifery and Palliative Care, King’s College London, UK; Department of Biostatistics & Health Informatics, IOPPN, King’s College London, UK; Department of Psychological Medicine, IOPPN, King’s College London, UK

## Abstract

**Background:** With rising referrals to child and adolescent mental health services (CAMHS), patient- and caregiver-facing digital platforms could streamline service delivery and research. With South London and Maudsley NHS Foundation Trust and King’s College London, the CAMHS Digital Lab developed myHealthE, a digital platform for families to access advice and support from the point of referral to CAMHS in South London, submit routine outcome measures, and consent to be contacted about relevant research studies.

**Method:** This secondary analysis descriptively summarised data from n=10,151 families under CAMHS invited to register on myHealthE between 2021 and 2023. Referral details, demographic and clinical characteristics were derived via the Clinical Record Interactive Search. myHealthE engagement was measured using Google Analytics. Routine outcome measures (particularly the Strengths and Difficulties Questionnaire [SDQ]) were collected via myHealthE.

**Results:** Of the invited families, 87.1% registered on myHealthE, and 85.4% completed at least one SDQ via the platform during their care episode. Overall, families completed n=29,906 SDQs during their episode of CAMHS support, mostly via myHealthE (93.2%) rather than conventional pen-and-paper methods (6.8%). Families engaged with resources on myHealthE, with 7,900 views of ‘Your Referral’ and ‘Information While You Wait’ pages over six-months. The platform also increased capacity for research recruitment, with 65.5% of families who consented for research contact over a 12-month period doing so via myHealthE.

**Conclusions:** myHealthE provides a valuable tool for patients, caregivers, clinicians and researchers. Such platforms can fulfil multiple functions. This is advantageous for both informing clinical service delivery, and for conducting research.

## Background

In 2024, 812,185 children and young people accessed mental health services in England (1). While the number of people in contact with Child and Adolescent Mental Health Services (CAMHS) has increased 410% since 2016, resources have struggled to keep pace with increasing demand; correspondingly, the number of doctors in child and adolescent psychiatry has only increased by 25% in the same period (2). Increasing pressure on services has resulted in long waiting times (3), during which young people and their families experience limited or no communication, causing considerable frustration and uncertainty (4).

Digital solutions could help overcome this strain on resources and increase timely access to support. Several patient-facing digital platforms have been developed and piloted in NHS CAMH services in England which can offer young people and families rapid access to advice and signposting. For example, the MyMind website, developed by Cheshire and Wirral Partnership NHS Foundation Trust with children and young people, provides practical information on mental health and CAMHS, and signposts to other sources of support and self-help resources, allowing users to select information based on their understanding of need (5). This platform was identified by the i-THRIVE Programme as a digital front-end example of best practice. Cam’s Den (Tavistock and Portman NHS Foundation Trust) (6), camhs (Humber NHS Foundation Trust) (7), and various other websites designed by local authorities and charities similarly offer mental health advice and signposting, but are not otherwise tailored to specific individuals or care pathways. Other platforms have been developed that fill this gap, in the form of virtual waiting rooms. The NCL Waiting Room, developed by the Tavistock and Portman NHS Foundation Trust, provides signposting and resources for children to develop self-care strategies while on the CAMHS waiting list (8). Children can also rate and track changes in their mood via the NCL Waiting Room, and communicate their needs and goals to therapists, although it is unclear whether this system addresses another crucial aspect of the patient journey: collecting routine outcome measures (ROMs) which integrate with CAMHS clinical systems.

Patient- and caregiver-rated ROMs help to assess intervention effectiveness for reducing symptoms, and track young people’s progress over an episode of care. ROMs are conventionally paper-based, but research has shown that ROMs are collected and logged in electronic health records for as few as 40% of patients, and of those patients, only 8% are followed up with subsequent ROMs (9). Collecting ROMs digitally could increase uptake. A number of tools have been developed to address this, although approaches to digital ROM collection are fragmented, and may not consistently integrate with other aspects of the patient journey (10). One example is POD, an online portal developed by the Anna Freud Centre for clinicians to gather ROMs from young people and caregivers (11). Scores are calculated and presented in tables and visualisations, supporting discussions about young people’s progress in-session, overcoming the delays and burdens associated with administering paper-based questionnaires and manual scoring. However, POD does not directly integrate into electronic health records, provide resources and signposting, or support recruitment to research studies which could complement treatment.

Overall, there is little evidence available on whether a CAMHS-oriented digital platform could enhance clinical care. This paper focuses on one such platform called myHealthE, developed by the CAMHS Digital Lab (12). The CAMHS Digital Lab implements digital innovations in CAMHS, and is based within the King’s Maudsley Partnership – a collaboration between King’s College London and South London and Maudsley NHS Foundation Trust (SLaM) (13). SLaM provides secondary mental health services, including CAMHS, to Croydon, Lambeth, Lewisham and Southwark, as well as some National & Specialist Services. In 2021, the CAMHS Digital Lab began a phased launch of myHealthE, which was developed with several aims:

- Ease pressure on clinicians to provide evidence-based psychoeducational material to young people and their families whilst they wait for direct clinical care;
- Help monitor ROMS as they change over time, and use this to inform patient care through automated integration into SLaM’s electronic health record;
- Provide patients with up-to-date information about their referral, and pathway-specific information as their assessment and treatment progresses;
- Support research delivery by providing data on symptoms and platform-use, and streamlining recruitment into research studies.

### myHealthE

This section provides an operational overview of myHealthE, from automated patient management to data collection and resource delivery (Figure 1; for more information on the displays, please see our introductory video) (14). Families are automatically invited to register on myHealthE following referral to SLaM CAMHS. An automated procedure identifies the cohort of families to be invited to myHealthE each week. Robotic Process Automation (RPA) enables direct entry into the myHealthE platform, facilitating enrolment of families. The data entered includes personal information and specifies which questionnaires each family is required to complete, along with the frequency of completion. The primary caregiver is then invited to register on the platform, where they can complete the specified ROMs. On this website, young people and caregivers can also access personalised resources in an ‘Information While You Wait’ and ‘About Your Service’ section, giving families interim support while they await clinical care. Families can also opt into research studies being undertaken in SLaM through the website (15, 16).

**Figure 1:**
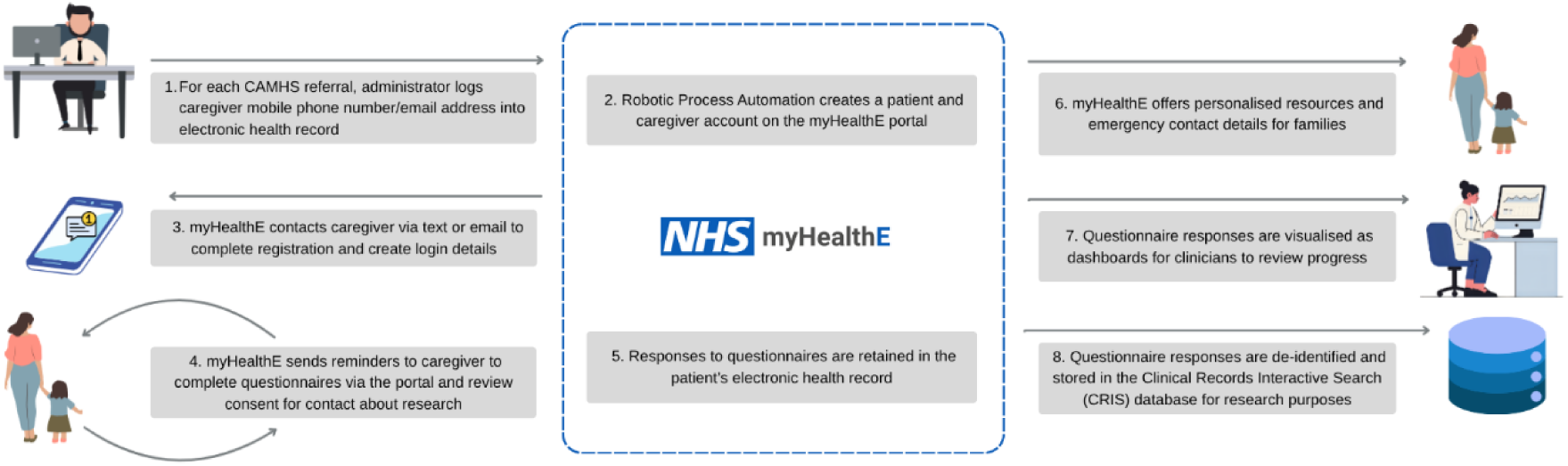
Operational overview of myHealthE.

ROMs collected through myHealthE are seamlessly integrated within the electronic Patient Journey System (ePJS), the electronic health record system used by clinicians in SLaM. myHealthE-collected ROM data are transformed into dynamic dashboards built using Microsoft Power BI, enabling clinicians to view ROMs for each young person, and for their caseload. The dashboards provide insights into ROM changes over time, as well as other key metrics. By enabling real-time ROM monitoring and delivering actionable feedback to both clinicians and families, the dashboards enhance clinical decision-making and promote data-driven, patient-centred care.

To ensure the data collected via myHealthE is available on ePJS for clinical use, we originally used a third-party company’s RPA system, and then developed our own approach to automate data entry through the front end of ePJS. To send families reminders of when their questionnaires are due, myHealthE incorporates two key components: a multi-agent tracking program which comprises the agents DataManager, UnitManager, and PatientFollowUp, as well as a supporting database (17). To meet hospital standards for handling patient-identifiable information, a comprehensive SLaM governance review ensured NHS Digital Technology Assessment Criteria compliance (18). The myHealthE system is hosted on a virtual server within SLaM’s Microsoft Azure cloud subscription. This configuration enables secure integration with SLaM’s IT infrastructure, including ePJS, and is protected by the same firewalls and cybersecurity measures as the rest of SLaM’s digital environment (17).

Data collected through myHealthE have significantly enhanced SLaM CAMHS’ research capabilities. Integrated within the Clinical Record Interactive Search (CRIS) system (19), ROMs collected via myHealthE contribute to de-identified datasets which are used to conduct research at scale. Researchers can therefore apply to access de-identified ROM data collected via myHealthE as part of other secondary data analyses conducted using CRIS.

### Stakeholder engagement

Co-production is at the heart of myHealthE’s design process. myHealthE was developed in collaboration with young people and caregivers, as well as the local community via the nonprofit organisation South London Listens. Regular stakeholder engagement has been instrumental in shaping myHealthE’s development; for instance, caregivers recommended visualising questionnaire responses, which was incorporated into the platform. The CAMHS Digital Lab team, who developed myHealthE, includes staff from King’s College London and SLaM, ensuring that clinicians and NHS informaticians are positioned as active decision-makers in our design process rather than as passive advisors. To incorporate co-design principles into the development process, we also collaborated with the Helen Hamlyn Institute’s Inclusive design group. Working with users at every stage of myHealthE development through ideation workshops, product testing, and more, has thus made it an accessible platform that meets families’ needs.

### Aims

The aim of this article is to provide an overview of the first families who were invited to engage with myHealthE since its launch in 2021, and provide illustrative examples of how myHealthE enhances both clinical delivery and CAMHS-oriented research.

## Methods

### Sample

We included all young people whose families were invited to register on myHealthE during the period 01/05/2021 to 31/12/2023. These families had been referred and accepted to SLaM’s community CAMHS, or to SLaM’s National & Specialist Service for Complex Autism and Associated Neurodevelopmental Disorders.

We defined a family’s episode of care as the accepted referral period during which they were invited to register on myHealthE. Invitation and registration to myHealthE may have taken place at any point during the episode of care. Some families had been discharged by the time of data extraction for this article; for others, their referrals were ongoing.

### Measures

We derived sociodemographic data for each young person from CRIS (19). Ethnicity (White, Black, Asian, Mixed, Other) (20) and income deprivation affecting children index (IDACI) national quintiles (21) were derived as categorical variables, although binary versions of ethnicity (White, All other ethnic groups) and IDACI (Most deprived, Least deprived) were used for analyses where cell counts were potentially disclosive. Similarly, sex was initially derived as a categorical variable (Female, Male, Other), but due to potentially disclosive cell counts, it could only be reported as a binary variable throughout (Female or Male, with Other treated as missing).

Clinical- and service-related factors were also extracted from CRIS (22). These included CAMHS locality, referral acceptance date, age at referral acceptance, assessment date (estimated as their first face-to-face or virtual attended event during the episode of care), treatment start date (estimated as their second face-to-face or virtual attended event during the episode of care), whether patients were referred to an ADHD treatment team, the earliest date of ADHD medication during the episode of care (derived using a natural language processing [NLP] tool, which uses artificial intelligence techniques to extract information from free-text notes), and whether patients had ever received various diagnoses.

For ROM data, we used the caregiver version of the Strengths and Difficulties Questionnaire (SDQ), completed either via myHealthE or via conventional methods. The SDQ is a brief emotional and behavioural screening questionnaire (23). It contains 25 items, with five items on each of five subscales: emotional problems, conduct problems, hyperactivity, peer problems, and prosocial behaviour. Users rate each item as Not True [0], Somewhat True [1], or Certainly True [2]. Some items are reverse-scored; scores are then summed across each subscale, and summed across the full scale to derive a total difficulties score. Cut-offs can be applied to indicate scores in normal, borderline and abnormal ranges (24) (although four-band cut-offs have also been derived, these three-band cut-offs are still used clinically; Supplement 1).

Meaningful change can also be derived based on movement between these three categories in a way which shows deterioration, improvement, or no change. The SDQ has shown satisfactory reliability and validity, with acceptable predictive validity for screening mental health disorders (23), although other studies have reported more mixed findings on its psychometric properties.

### Statistical analysis

We primarily report descriptive statistics for categorical variables (frequencies, percentages) and continuous variables (median, interquartile range [IQR]). Where relevant, we also report results of Chi-square tests. To provide a geographical perspective on who contributes ROM data via myHealthE, heat maps were produced using 2011 Lower Super Output Area (LSOA) boundary data retrieved from the UK Data Service (25). Analyses were conducted in Stata (version 18.0), and data visualisation in R (version 4.2.1).

## Results

### Sample profile

Of the n=10,151 families referred to CAMHS and invited to register on myHealthE, the majority went on to register (n=8,842, 87.1%) and complete at least one SDQ via the platform during their episode of care (n=8,673, 85.4%) (Figure 1). In total, n=8,881 (87.5%) completed at least one SDQ using any method between referral acceptance and discharge/data extraction. The total number of SDQs completed across these families during their episode of care was n=29,906, ranging from 1 to 18 per young person (median = 3, IQR = 1 to 5). The majority of these SDQs were completed via myHealthE (n=27,862, 93.2%), rather than by conventional methods (n=2,044, 6.8%).

Sample characteristics are summarised in Table 1. We also show sample characteristics for families who only ever completed an SDQ via conventional methods during their episode of care, to demonstrate that relying on SDQs completed through this route results in a much smaller sample, categories which are too small to use in complex statistical analysis, and distributions of sample characteristics which are not representative of the overall cohort.

**Table 1:** Sample characteristics.

|  | Invited to<br>myHealthE<br>(n=10,151) | Completed SDQ via<br>conventional<br>methods only during<br>care episode<br>(n=208) | Completed SDQ via<br>conventional methods or<br>myHealthE during care<br>episode<br>(n=8,881) |
| --- | --- | --- | --- |
| **Sex (n, %)** |  |  |  |
| Female | 4,441 (44.0%) | 118 (57.6%) | 3,800 (43.2%) |
| Male | 5,652 (56.0%) | 87 (42.4%) | 5,033 (57.0%) |
| **Ethnicity (n, %)** |  |  |  |
| White | 4,671 (46.9%) | 93 (45.6%) | 4,180 (48.0%) |
| Black | 2,252 (22.6%) | 45 (22.1%) | 1,902 (21.8%) |
| Asian | 474 (4.8%) | 11 (5.4%) | 398 (4.6%) |
| Mixed | 1,878 (18.9%) | 40 (19.6%) | 1,662 (19.1%) |
| Other | 686 (6.9%) | 15 (7.4%) | 576 (6.6%) |
| **IDACI (n, %)** |  |  |  |
| Most deprived | 7,889 (79.9%) | 169 (82.4%) | 6,883 (79.5%) |
| Least deprived | 1,991 (20.2%) | 36 (17.6%) | 1,772 (20.5%) |
| **Service borough (n, %)** |  |  |  |
| Croydon | 3,156 (31.2%) | 20 (9.7%) | 2,855 (32.2%) |
| Lambeth | 1,793 (17.7%) | 65 (31.4%) | 1,563 (17.6%) |
| Lewisham | 2,549 (25.2%) | 61 (29.5%) | 2,201 (24.8%) |
| Southwark | 2,361 (23.3%) | 48 (23.2%) | 2,022 (22.8%) |
| National & Specialist | 270 (2.7%) | 13 (6.3%) | 223 (2.5%) |
| **Age at referral**<br>**acceptance in years**<br>**(median, IQR)** | 11 (8 to 13) | 12 (10 to 14) | 10 (7 to 13) |

Notes: Overall, sex was missing or 'Other' for n=58 (0.6%), ethnicity was missing for n=190 (1.9%), IDACI was missing for n=271 (2.7%), borough was missing for n=22 (0.2%), and age at referral acceptance was missing for n=2 (<0.1%).

Supplementing with SDQs completed via the myHealthE platform overcomes these biases, approximating the underlying cohort much more closely. Figure 2 also illustrates that, among families resident in the local catchment area and discharged by the time of data extraction, the number of SDQs they completed during their episode of care did not appear to vary substantially according to area-level deprivation.

**Figure 2:**
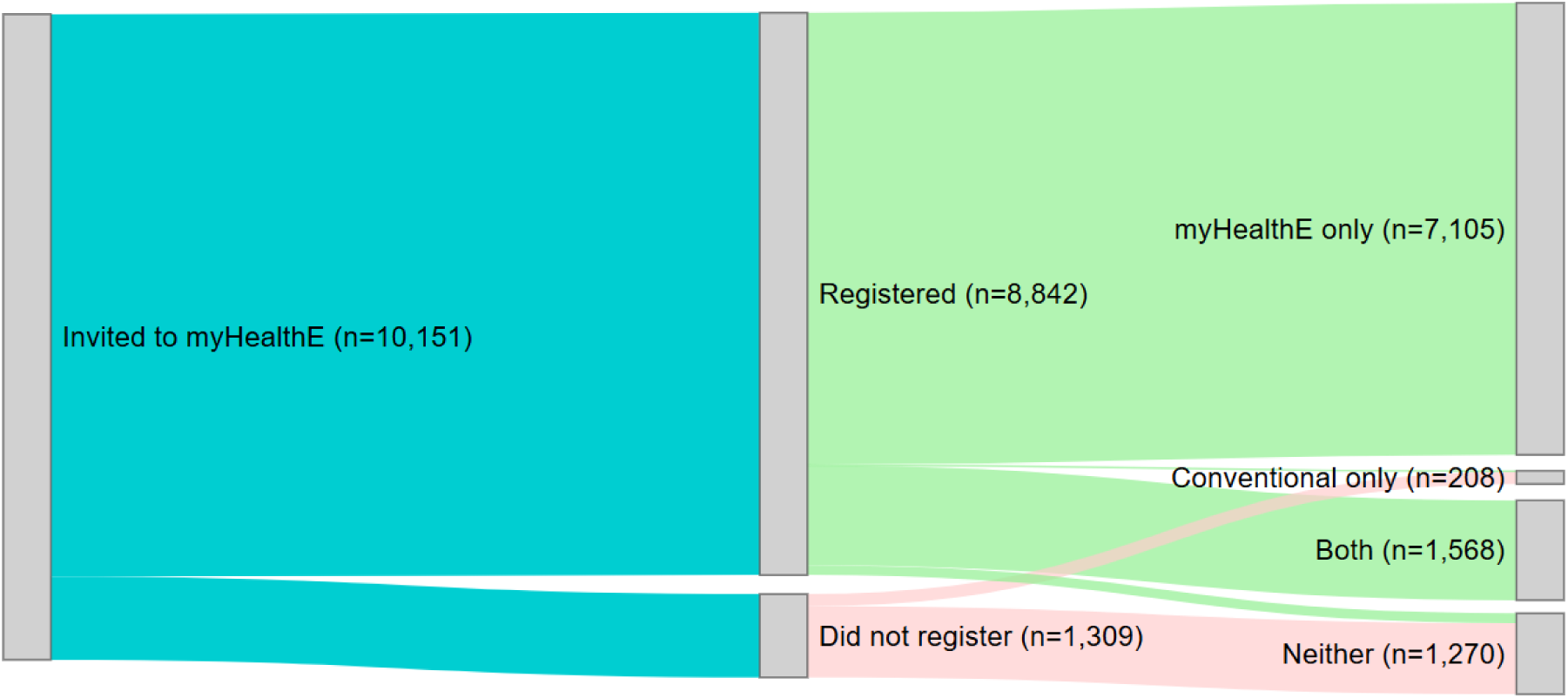
Sankey diagram showing the number of families referred to CAMHS and invited to register on myHealthE (left), the number who registered on myHealthE (middle), and the formats in which they completed SDQs (right).

**Figure 3:**
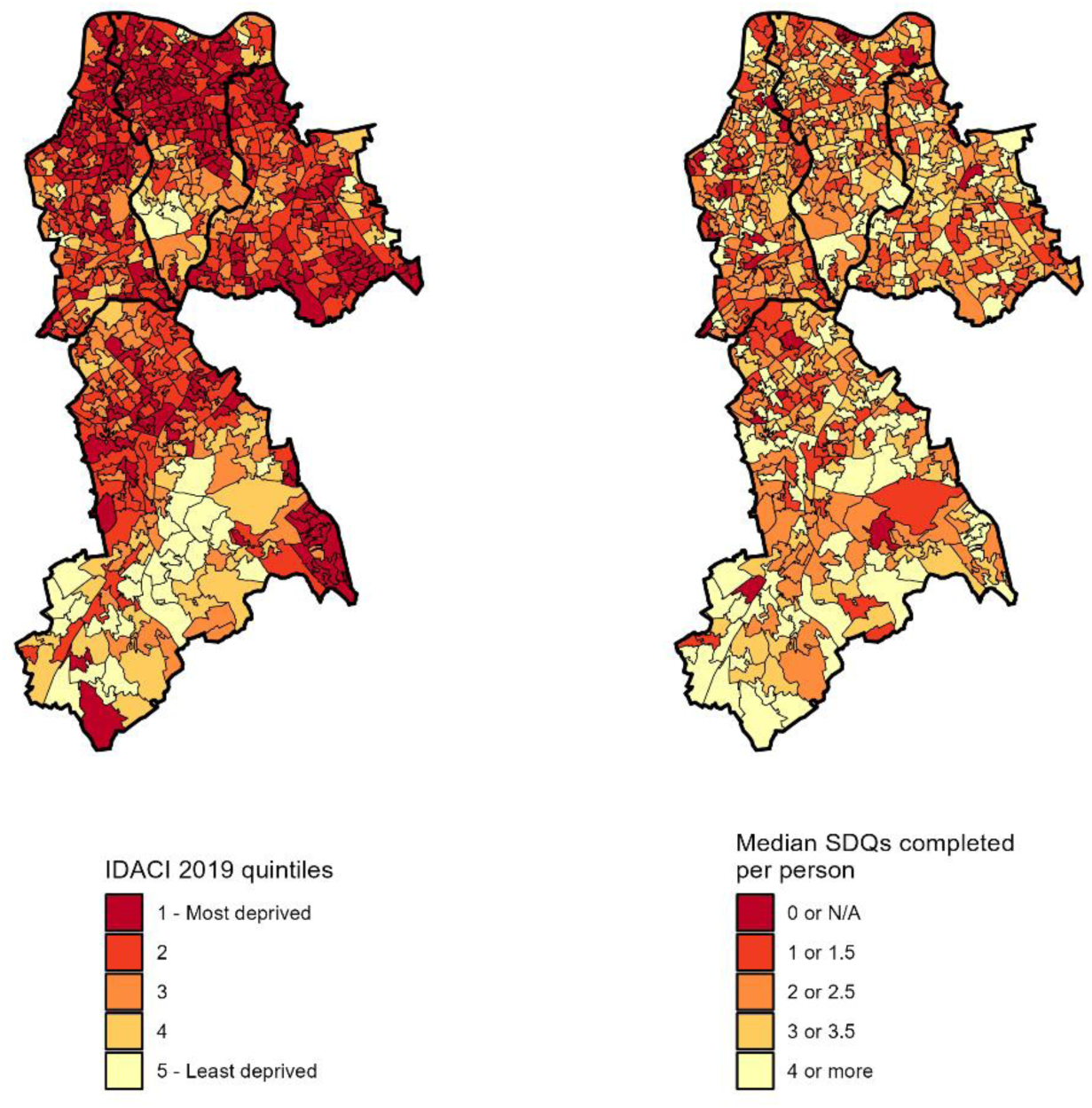
Area-level deprivation (left) and median SDQs completed per person during episode of care (right) by Lower Super Output Area.

### Illustrative research and clinical applications

In this section, we provide examples of how increased ROM availability through myHealthE supports clinical delivery and health service research throughout CAMHS’ pathways.

### Referral and triage

#### Clinical application

Several SLaM CAMHS single point of access teams (26) made use of ROM data during the triage process to review young people’s symptoms and inform the type of assessment and care they should receive. For example, in our sample, n=384 young people were referred to an ADHD pathway after being invited to myHealthE, of whom n=242 completed an SDQ prior to treatment commencement. As would be expected, the median score on the SDQ’s hyperactivity subscale in this group was in the abnormal range (9.5, IQR = 8 to 10, abnormal range = 7 to 10). Some teams used high hyperactivity scores at referral to then trigger more specific ADHD-related measures, including further review of symptoms through more detailed questionnaires (such as the SNAP [Swanson, Nolan, and Pelham] rating scale), which were also delivered through myHealthE. Feedback from the services described this as a more efficient method to inform plans for subsequent CAMHS care.

#### Research application

Researchers have begun to capitalise on these advances in data acquisition. For example, within our clinical services, research is underway to examine sociodemographic differences in ADHD symptoms and subsequent treatment pathways, with a focus on sociodemographic differences in presentation with hyperactivity symptoms (27).

Preliminary data show that a small proportion of young people under ADHD treatment teams had hyperactivity scores in the normal (n=17, 7.0%) or borderline (n=16, 6.6%) range. Those referred to ADHD treatment teams despite having normal or borderline SDQ hyperactivity scores were less frequently living in deprived areas (Table 2).

**Table 2:** SDQ hyperactivity scores among patients referred to ADHD treatment teams, stratified by sociodemographic characteristics.

|  |  | Normal/borderline<br>hyperactivity SDQ<br>score (n=33) | Abnormal<br>hyperactivity SDQ score<br>(n=209) |
| --- | --- | --- | --- |
| **Sex (n, %)** |  |  |  |
|  | *Female* | 10 (30.3%) | 56 (26.8%) |
|  | *Male* | 23 (69.7%) | 153 (73.2%) |
| **Ethnicity (n, %)** |  |  |  |
|  | *White* | 16 (48.5%) | 117 (56.3%) |
|  | *All other ethnic groups* | 17 (51.5%) | 91 (43.8%) |
| **IDACI (n, %)** |  |  |  |
|  | *Most deprived* | 22 (66.7%) | 156 (75.7%) |
|  | *Least deprived* | 11 (33.3%) | 50 (24.3%) |
| **Age at referral**<br>**acceptance in years**<br>**(median, IQR)** |  | 11 (9 to 13) | 10 (7 to 12) |

Notes: Ethnicity was missing for n=1 (0.4%), and IDACI for n=3 (1.2%).

### Waiting period

#### Clinical application

In the final six months of this study alone (July to December 2023), there were 3,171 views of the ‘Information While You Wait’ page on myHealthE, and 4,729 views of the ‘About Your Service’ page, according to Google Analytics (7,900 views in total). We are examining how services can use ROM data submitted during the waiting period to personalise these resources and signpost to other services, tools, and sources of support while awaiting assessment (16). In our sample, n=6,665 (65.7%) families completed an SDQ sometime between referral acceptance and assessment. Symptoms reported in the first such SDQ during this period showed a diverse range of problems faced by these young people (Table 3) – resources presented to them in the virtual waiting room could be tailored accordingly. Of the young people who had undergone assessment by the time of data extraction, n=3,065 had completed more than one SDQ during the waiting period – for these young people, clinicians can therefore also see how their SDQ scores change, highlighting those who might be deteriorating and in need of more urgent care (Table 3).

**Table 3:**
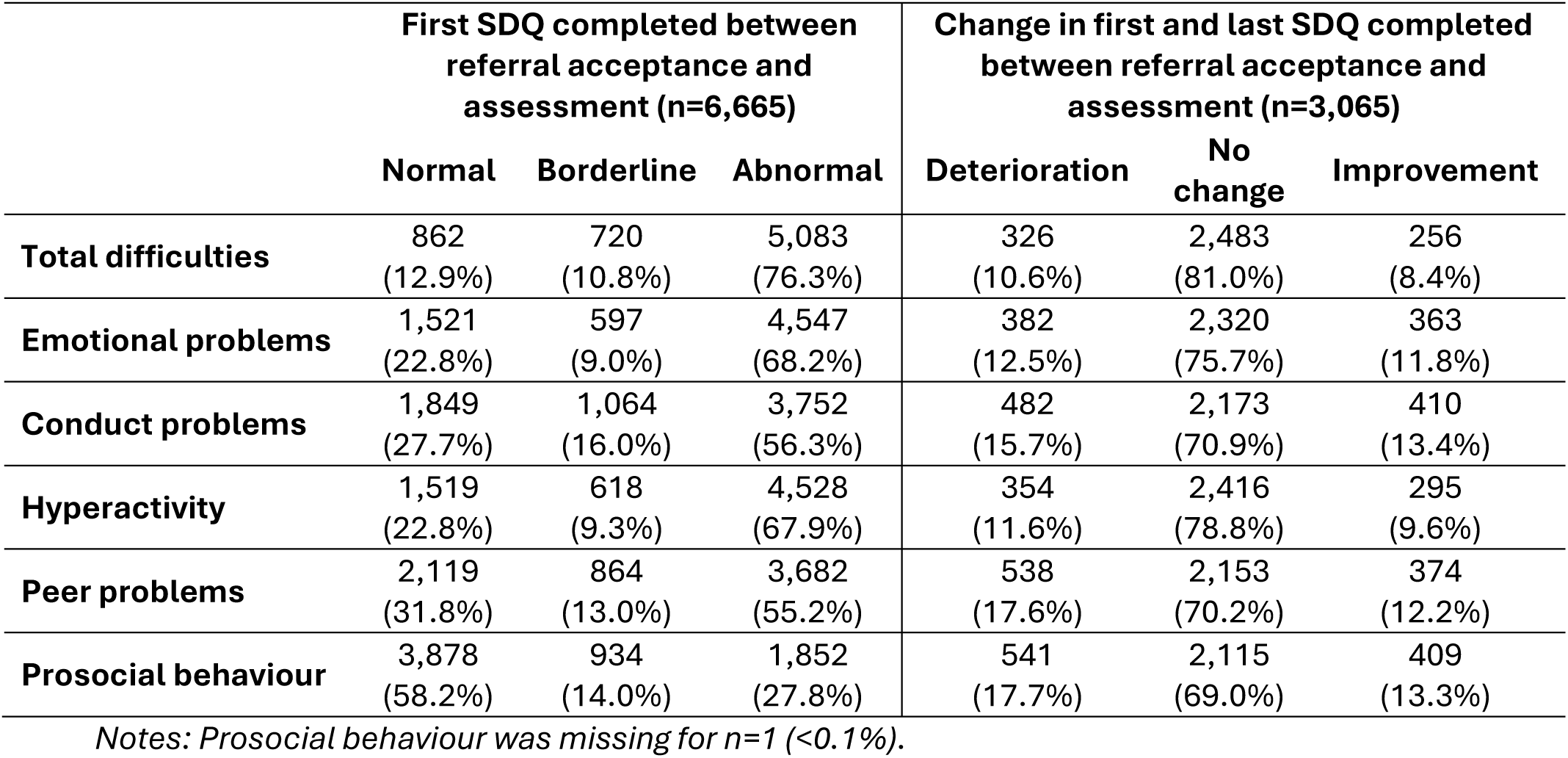
SDQs completed between referral acceptance and assessment.

|  | First SDQ completed between referral acceptance and assessment (n=6,665) |  |  | Change in first and last SDQ completed between referral acceptance and assessment (n=3,065) |  |  |
| --- | --- | --- | --- | --- | --- | --- |
|  | Normal | Borderline | Abnormal | Deterioration | No change | Improvement |
| **Total difficulties** | 862<br>(12.9%) | 720<br>(10.8%) | 5,083<br>(76.3%) | 326<br>(10.6%) | 2,483<br>(81.0%) | 256<br>(8.4%) |
| **Emotional problems** | 1,521<br>(22.8%) | 597<br>(9.0%) | 4,547<br>(68.2%) | 382<br>(12.5%) | 2,320<br>(75.7%) | 363<br>(11.8%) |
| **Conduct problems** | 1,849<br>(27.7%) | 1,064<br>(16.0%) | 3,752<br>(56.3%) | 482<br>(15.7%) | 2,173<br>(70.9%) | 410<br>(13.4%) |
| **Hyperactivity** | 1,519<br>(22.8%) | 618<br>(9.3%) | 4,528<br>(67.9%) | 354<br>(11.6%) | 2,416<br>(78.8%) | 295<br>(9.6%) |
| **Peer problems** | 2,119<br>(31.8%) | 864<br>(13.0%) | 3,682<br>(55.2%) | 538<br>(17.6%) | 2,153<br>(70.2%) | 374<br>(12.2%) |
| **Prosocial behaviour** | 3,878<br>(58.2%) | 934<br>(14.0%) | 1,852<br>(27.8%) | 541<br>(17.7%) | 2,115<br>(69.0%) | 409<br>(13.3%) |

*Notes: Prosocial behaviour was missing for n=1 (<0.1%).*

#### Research application

Research is investigating potential biases or mechanisms underlying these symptom changes while awaiting assessment. Initial explorations show associations between young people’s sex, changes in emotional problems (X^2^[2]=33.0, p<0.001), and changes in hyperactivity (X^2^[2]=29.9, p<0.001) (Figure 4) (no statistically significant associations were observed between sex and changes on other subscales). Tentatively, females more frequently show stability in their emotional symptoms during the waiting period, while males more frequently show stability in their hyperactivity symptoms.

**Figure 4:**
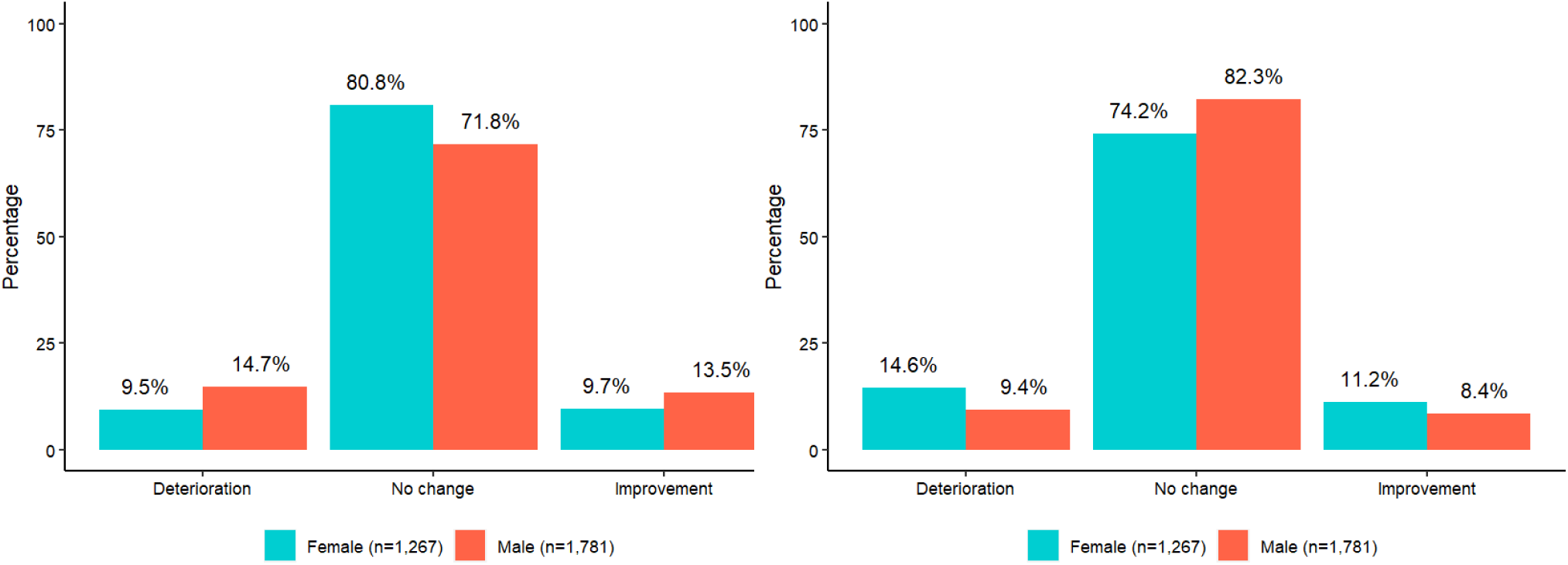
Changes in emotional problems (left) and hyperactivity (right) during the waiting period between referral acceptance and assessment, stratified by sex.

Information on young people’s symptoms, and how they change while on the waiting list, could be informative for trial recruitment during this period by highlighting families who meet eligibility criteria for experimental studies. Indeed, feasibility work has shown early promise for using myHealthE as a waitlist recruitment tool (16), and there are plans to incorporate randomisation functionalities into the platform. Recently, a ‘consent for contact’ function was enabled in myHealthE, so that families can consent to be contacted about research opportunities through the platform. From 1^st^ February 2024 to 31^st^ January 2025, 65.5% of the n=1,702 families who consented for research contact did so via myHealthE, illustrating its potential as a research recruitment tool.

### Assessment

#### Clinical application

At the point of assessment, clinicians review ROMs that families have submitted over the waiting period via dashboards to inform assessment and support diagnosis. This has proved helpful to clinicians for tracking changes in problem severity and priority since their initial referral to CAMHS. For example, among the families in our cohort who completed multiple SDQs during the waiting period, n=266 young people scored in the abnormal range for emotional problems on their first SDQ, but no longer scored in the abnormal range in their most recent SDQ to assessment. Instead, some of these young people scored in the abnormal range for conduct problems (n=18), hyperactivity (n=16), and peer problems (n=19) where they had not done previously. This is helping to inform the care pathway young people join following assessment, particularly with the addition of other ROMs to the myHealthE platform, such as the Revised Children’s Anxiety and Depression Scale (RCADS), further supporting assessment and diagnosis.

#### Research application

Research is underway to investigate the predictive validity of ROMs for different clinical diagnoses. Initial exploration of the first SDQs completed by our cohort indicated varying subscale scores across different diagnostic groups (Table 4). Predictive models are being developed to understand how ROMs completed during the waiting period can accurately predict certain diagnoses, whilst taking into account other information like sociodemographic characteristics (28).

**Table 4:**
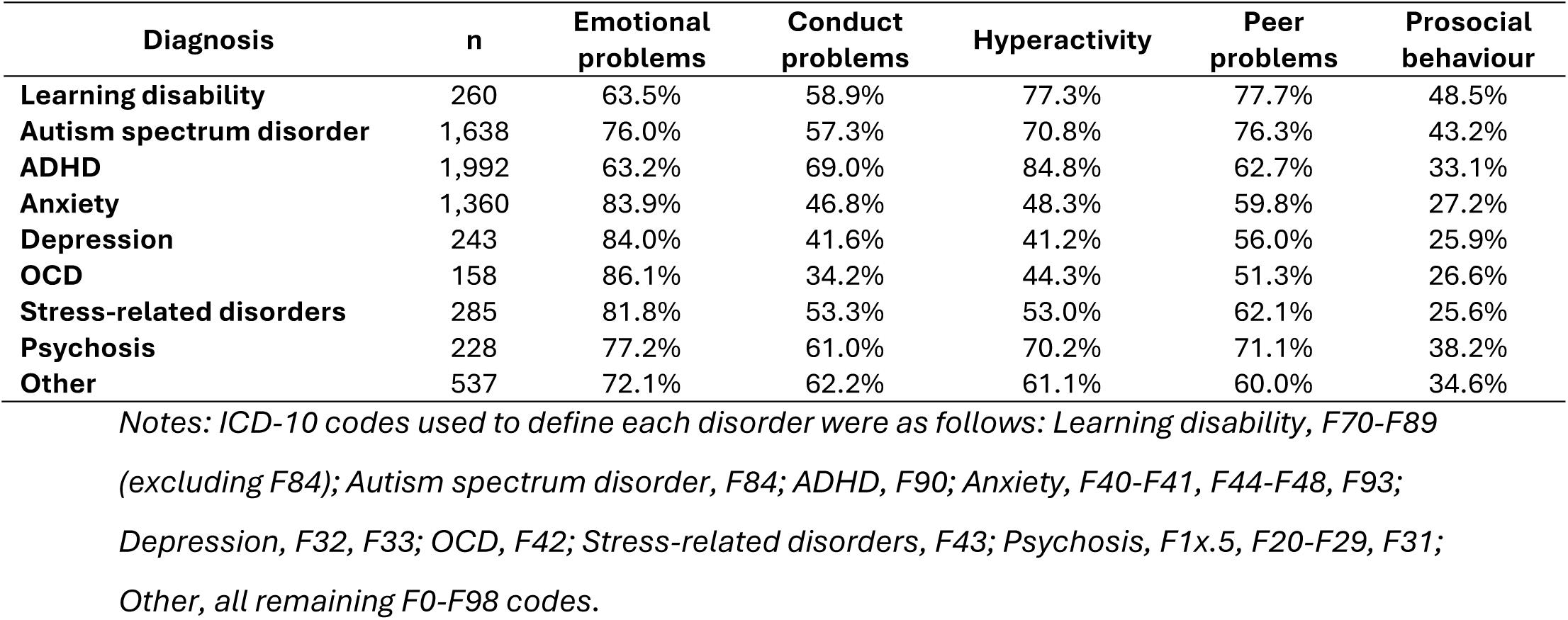
Proportion of young people ever diagnosed with each disorder who scored in the abnormal range for each SDQ subscale.

| Diagnosis | n | Emotional problems | Conduct problems | Hyperactivity | Peer problems | Prosocial behaviour |
| --- | --- | --- | --- | --- | --- | --- |
| Learning disability | 260 | 63.5% | 58.9% | 77.3% | 77.7% | 48.5% |
| Autism spectrum disorder | 1,638 | 76.0% | 57.3% | 70.8% | 76.3% | 43.2% |
| ADHD | 1,992 | 63.2% | 69.0% | 84.8% | 62.7% | 33.1% |
| Anxiety | 1,360 | 83.9% | 46.8% | 48.3% | 59.8% | 27.2% |
| Depression | 243 | 84.0% | 41.6% | 41.2% | 56.0% | 25.9% |
| OCD | 158 | 86.1% | 34.2% | 44.3% | 51.3% | 26.6% |
| Stress-related disorders | 285 | 81.8% | 53.3% | 53.0% | 62.1% | 25.6% |
| Psychosis | 228 | 77.2% | 61.0% | 70.2% | 71.1% | 38.2% |
| Other | 537 | 72.1% | 62.2% | 61.1% | 60.0% | 34.6% |

*Notes: ICD-10 codes used to define each disorder were as follows: Learning disability, F70-F89 (excluding F84); Autism spectrum disorder, F84; ADHD, F90; Anxiety, F40-F41, F44-F48, F93; Depression, F32, F33; OCD, F42; Stress-related disorders, F43; Psychosis, F1x.5, F20-F29, F31; Other, all remaining F0-F98 codes.*

### Treatment

#### Clinical application

Clinicians can review ROMs during treatment to monitor changing symptoms and treatment response. Figure 5 shows changing SDQ total difficulties scores for n=985 families who had completed at least two SDQs in the period between treatment commencement and discharge. While some showed improvement over the period (n=151, 15.3%), others showed either no change (n=753, 76.5%) or deterioration (n=81, 8.2%). For young people who show no improvement, clinicians might consider alternative treatment approaches.

**Figure 5:**
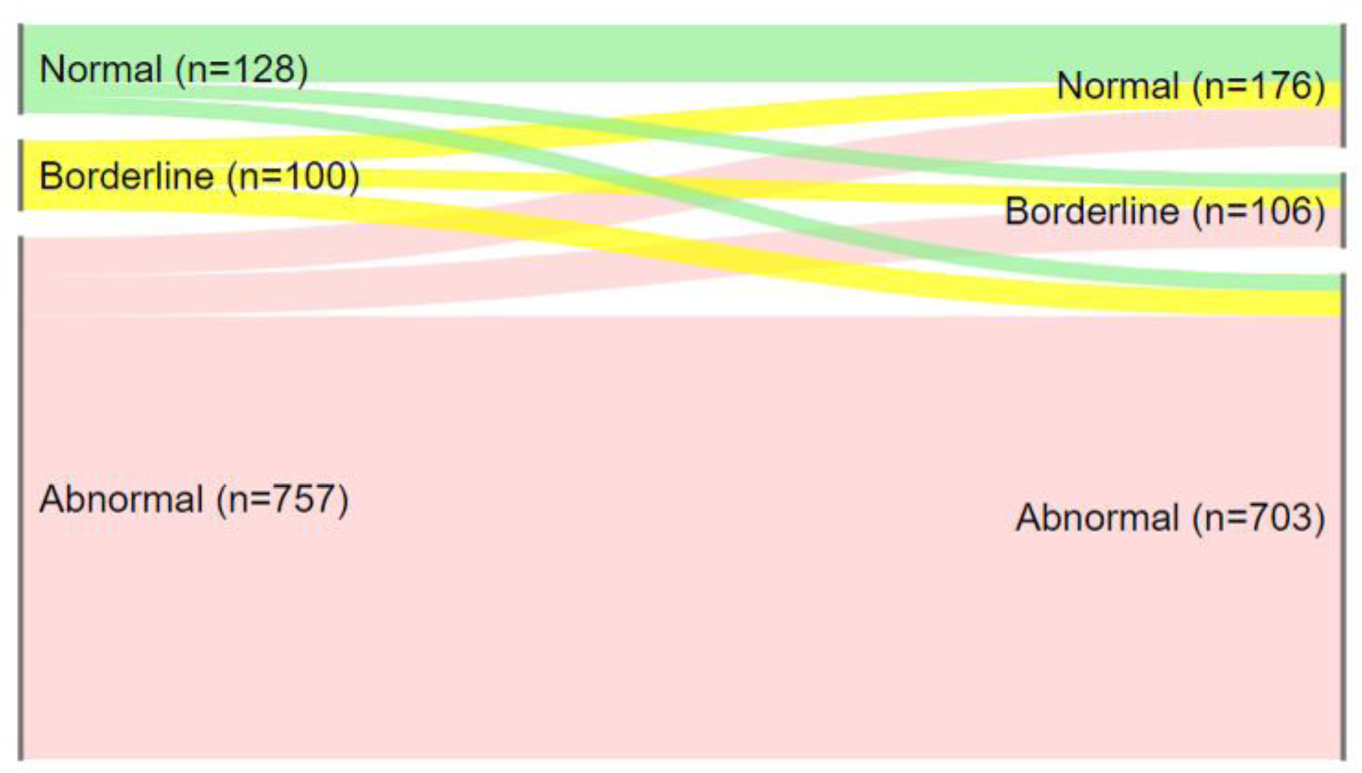
Changing SDQ total difficulties scores between the first and last SDQs completed in the period between treatment commencement and discharge.

#### Research application

Studies are underway which use ROM data combined with information derived using NLP tools to investigate which treatments work for whom. For example, in our cohort, n=368 young people started ADHD medication sometime during their episode of care (identified using NLP), and fully completed at least one SDQ both before and after the first mention of ADHD medication in their clinical notes. Of this group, n=176 (47.8%) showed improvement on any SDQ subscale before and after this first medication mention. Quasi-experimental studies or target trial emulation could be conducted using these data to compare the effectiveness of different ADHD treatment approaches, potentially drawing outcomes from other data sources as well, such as actigraphy data from wearables (currently being developed as part of the Paediatric Actigraphic Clinical Evaluation project) (29).

### Discharge

#### Clinical application

Clinicians are exploring methods of staged discharge using myHealthE as a digital resource, where young people and families are offered tailored digital resources to sustain recovery post-discharge. If myHealthE usage can be sustained post-discharge, this could also be informative for clinicians if a subsequent referral is made. For example, for young people navigating the transition from CAMHS to adult mental health services, clinicians in adult mental health services could review changing symptoms during any transitional periods where young people may not be receiving direct care to inform further assessment and care planning. In our sample, n=314 families completed further SDQs via myHealthE after being discharged.

Caregivers have reported finding the longitudinal monitoring data helpful, as well as the supportive resources (30).

The platform is also being used to rapidly roll out surveys for families to provide feedback on their experiences of accessing CAMHS. One CAMHS locality is piloting an adapted feature of myHealthE to collect de-identified patient experience questionnaires. So far, this has substantially boosted survey response rates – preliminary analysis indicates that median survey responses per month increased from 13 to 122, an 838% increase. This shows promise for effective service evaluation and planning.

#### Research application

Continuing ROM collection after discharge could drive research on the recovery and remission of young people after accessing CAMHS. This information could be supplemented using data from external sources. For example, CRIS records for CAMHS patients have been linked to the National Pupil Database, which contains education records for pupils in England’s state schools (22). Using these data, we could investigate the long-term educational outcomes of young people who have been discharged from CAMHS, including whether any lasting changes in mental health symptoms post-discharge are accompanied by improvements in functional outcomes such as educational attainment or school absence.

## Discussion

myHealthE is the first platform of its kind to be fully integrated into NHS CAMHS. Previous platforms have been developed that variously provide resources, virtual waiting rooms, and support ROM collection for CAMHS populations. This is the first such platform to achieve all three, providing a unifying ‘one-stop shop’ for young people and caregivers, and the first to be meaningfully integrated into clinical practice and electronic health record systems. By providing an overview of available ROM data following myHealthE’s first two years of implementation, we have demonstrated that it has the functionality to inform every stage of the patient care pathway through CAMHS, as a tool for clinicians, families, and researchers.

myHealthE’s resources and virtual waiting room support young people and caregivers, particularly while they await assessment and treatment from CAMHS, hopefully increasing transparency and mitigating some of the uncertainty and frustration associated with long waiting times (4). This also reduces the burden on clinicians to identify and disseminate relevant resources to individual families. Higher ROM completion rates through the platform also allow us to track symptom changes during care episodes more closely. Presented back to clinicians through dashboards, this information can inform treatment approaches, and may in future help us to tailor the resources shown to families in the virtual waiting room, leading to more personalised support.

From a research perspective, higher ROM completion rates allow us to conduct service evaluation and research to inform service delivery with greater statistical power and reduced response bias. This is evidenced by the higher cell sizes and greater representativeness of the underlying sample when drawing ROM data from myHealthE. Used in combination with our wider research infrastructure, including CRIS (19), NLP tools (31), and external data linkages (22), this increases our capacity to conduct impactful, rigorous, and cross-disciplinary research in the field of child and adolescent mental health. Over the coming years, we will be continuing our research in this area leveraging these data.

Many barriers were encountered when implementing myHealthE into routine NHS services, such as navigating information governance requirements, existing IT infrastructure, and NHS networks and firewalls. These challenges are detailed elsewhere (17). In future, we hope to collaborate with other NHS Trusts and third sector organisations to expand myHealthE into other services offering child and adolescent mental health support. This expansion will necessitate overcoming similar barriers in other settings, but myHealthE’s successful implementation in SLaM provides a model for how this might be achieved in these sites.

While these operational challenges have been overcome in our local CAMHS, other limitations remain. Digital exclusion, whereby groups disengage with technology either out of personal choice or for reasons beyond their control, remains a concern for policymakers (32). Research on the digital divide has highlighted that moves towards digital platforms may particularly disadvantage some ethnic, socioeconomic, age and language groups (33, 34). Our sample did not show any strong digital divides, particularly as compared to biases that may arise from relying on conventionally applied pen-and-paper ROMs. Nonetheless, we plan to continue work to make myHealthE as accessible as possible, for example through translation to other languages. Inequalities in service access also remain a wider issue in CAMHS (35), so while those who submit ROMs via our platform might be broadly representative of CAMHS patients, they are not necessarily representative of children and young people in the wider population who have mental health difficulties. Other areas for improving myHealthE also remain, for example enabling ROM submission by multiple caregivers (rather than just the primary caregiver), and enabling self-report ROM submissions by young people. The addition of further ROMs will also be an important aspect of myHealthE’s development, considering mixed findings on the SDQ’s psychometric properties and therefore its utility as an outcome measure for clinicians and researchers alike (36).

Nonetheless, myHealthE presents a valuable tool for young people, caregivers, clinicians and researchers. Patient-facing platforms can fulfil multiple functions, like offering a virtual waiting room space, providing advice and resources, supporting ROM collection, and boosting research recruitment. These platforms are especially valuable if they can be integrated into existing clinical record systems, and wider research infrastructure.

## Data Availability

The data cannot be made publicly available, but can be accessed with permissions from South London and Maudsley NHS Foundation Trust.

## Acknowledgements

At the time of conducting this work, the CAMHS Digital Lab comprised: AW, CC, SRB, JH, JA, RS, SJL, AM, ES, LZ, JP, JD, Zoë Firth, Stephen Douch, Sophie Epstein, Shuo Zhang, Laurence Telesia, David Howard, Senta Haeussler, Brian Ching, Isabel Yorke, Nicholas Cummins, Judith Dineley, Asilay Seker, Akash Roy Choudhury, Garry Moriarty, Daniel Smith, Sam Harris, Cato Zantman, Su Mon Latt, Nicoletta Adamo. We are grateful for ongoing support from clinical and academic colleagues within the King’s Maudsley Partnership and beyond - Harold Bennison, Bruce Clark, Adam Shortland, Zina Ibrahim, Stuart Maclellan, Edmund Sonuga-Barke, Jacqueline Phillips-Owens, Sarah Holloway, Sara Saunders, Nicola Dykes, Matthew Hotopf, Robert Stewart, Richard Dobson, Matthew Broadbent, Claire Delaney-Pope, Jayati Das-Munshi, Omer Moghraby, Anto Ingrassia, Amanda Waldman, Manish Rao and many more.

Map data is provided with the support of the ESRC and JISC and uses boundary material which is copyright of the Crown, the Post Office and the ED-LINE consortium. Contains National Statistics data © Crown copyright and database right 2024. Contains OS data © Crown copyright and database right 2024.

## Author Contributions

All named authors contributed to the manuscript drafting and/or review. All authors approve the manuscript for submission.

