## Supplement 1 for "Scaling a digital platform for informing and improving child and adolescent mental health service delivery and research: results from the first 10,000 families engaged with myHealthE"

*Supplement 1: Caregiver SDQ cut-offs*

| SDQ subscale | Normal | Borderline | Abnormal |
|---|---|---|---|
| Total difficulties | 0-13 | 14-16 | 17-40 |
| Emotional problems | 0-3 | 4 | 5-10 |
| Conduct problems | 0-2 | 3 | 4-10 |
| Hyperactivity | 0-5 | 6 | 7-10 |
| Peer problems | 0-2 | 3 | 4-10 |
| Prosocial behaviour | 6-10 | 5 | 0-4 |

Source: https://www.sdqinfo.org/py/sdqinfo/c0.py
